# Bariatric surgery: preparations and quality of life consequences

**DOI:** 10.1101/2022.12.29.22283988

**Authors:** Liis Lozano, Triin Põder, George A. Lozano

## Abstract

**Introduction:** Obesity is a major threat to global health. When more conventional methods have failed, obesity can be addressed via bariatric surgery. Here we examine the reasons why patients choose bariatric surgery and the behavioral consequences thereafter.

**Methods:** A qualitative study with a phenomenological design was used to analyze detailed interview responses from recipients of bariatric surgery.

**Results:** Before surgery, (a) bariatric surgery was chosen for obvious reasons: patients had been overweight, had obesity-related health problems, and had difficulties moving. (b) Information was obtained from medical practitioners and online discussion groups. (c) Fear, anxiety, and apprehension were common. Friends and family were sometimes supportive and sometimes disapproving. After surgery, (1) many chronic health problems disappeared or eased significantly. (2) Subjects had difficulties adjusting to small portions and altered food preferences. (3) Physical activity increased. (4) Self-esteem increased but the worry of regaining the weight remained. (5) As before surgery, there were both supportive and condemning attitudes by relatives, friends, and society. (6) A desire for further nutritional and psychological counseling was indicated. (7) Relationships and quality of sex improved in most cases but not always. Single women, particularly, became more active in potential relationships.

**Discussion:** With a few exceptions, our results agree with the literature, supporting the idea that bariatric surgery leads to extensive physical, psychological, and social changes. Hence, patients ought to be better prepared for these changes, and medical practitioners ought to be aware of the magnitude of the changes this surgery will bring about in their patients’ lives.

## INTRODUCTION

Over the past fifty years, obesity has become a major global health concern^1,2^. Obesity decreases life expectancy and quality of life by increasing the risk and severity of many diseases, among them type-2 diabetes^3^, cardiovascular disease^4^, asthma^5^, covid-19^6^, sleep apnoea^7^, periodontal disease^8^, hypertension^9^, osteoarthritis, fatty liver disease, stroke, dementia, and several cancers^1,10^. Although obesity as an epidemiological concern began in the world’s most affluent counties^2,11^, as the global standard of living has risen, obesity has also become a problem in developing countries^12,13^. As of 2020, over 1.9 billion people in the world were overweight, of which over 650 million were obese^14^.

Many approaches have been used to combat obesity^15,16^. Epidemiologically, programs that increase the activity level^17^ or alter people’s diet^18^ have been implemented. In addition, several evolutionary explanations for obesity have been proposed, each one with its own prognostic solutions^19-21^. At the individual level, obesity can be addressed by several means, among them diet modification^22,23^, increased activity^24,25^, cognitive therapy^26,27^, and appetite suppressive drugs^28,29^. When other options have failed, bariatric surgery becomes a viable alternative^30^.

Bariatric surgery is a laparoscopic procedure whereby the volume of the stomach and/or the length of the small intestine are reduced^31,32^. There are several types of surgeries, affecting the size of the stomach, the path food takes, and the amount and section of intestine removed. Bariatric surgery has two general effects: first, the amount of food that can be consumed at a given feeding bout is reduced, and second, the absorption of this food also decreases^33,34^. Bariatric surgery is one of the few effective long-term cures for severe obesity, and it also relieves many associated conditions, leading to an increase of both lifespan and quality of life^30,35^.

As with any surgery, bariatric surgery patients receive pre-op information and post-op care instructions. However, unlike some other surgical procedures, bariatric surgery leads not only physical changes, but also social and psychological changes^36^. Information necessary to help patients deal with these major changes in their lives is relatively recent and still limited^37^. In Estonia, there has been work on the preparation necessary for surgery and post-surgical complications^38^ and on post-surgical nursing-care recommendations^39^. However, no research exists on the social and psychological effects. Hence, the aim of our research is to examine from a psychological, social, and medical perspective: (1) the preparations for bariatric surgery, and (2) the consequences after surgery.

## METHODS

### Background information

The research was conducted in Estonia, where as of 2016, 35.2% of the population was overweight (BMI > 25) and 19.2% obese (BMI >30)^40^. The costs of bariatric surgery in Estonia are covered by the government for people with a BMI of over 40 who have not been able to lose weight by other means, and for people with a BMI of over 35 who have at least one obesity-related disease^41^.

From 2004 to 2017, 5752 bariatric surgeries were performed in Estonia. In the first couple of years, fewer than 10 surgeries per year were conducted but the frequency has been increasing, and since 2013, the number of surgeries has been stable at almost 700 surgeries per year. In 2017, there were 560 surgeries per 1 million people, a rate that is similar to that of Norway (566) and Sweden (565), and higher than the rate in Finland (166) and in Denmark (125)^42^.

### Study subjects

Participants were recruited using an Estonian Facebook social media group called “Patients of bariatric surgery”, created on July 2014 and as of January 21, 2022, had 974 members. An invitation was sent to the group. Interested people were told about the aims of the research, and those who agreed to participate were asked to read and sign an informed consent form. Participants were adults (≥ 18 years old) who had undergone bariatric surgery at least a year before.

### Interviews

This research is qualitative and phenomenological. Qualitative methods of research address everything that is connected with the issue at hand: people’s experiences, feelings, behavior, expectations, and beliefs. Phenomenological research methods examine a person’s experience, usually by means of an interview^43^.

Participants were asked three questions related to their experiences before surgery (Tab. 1) and seven related to their experiences after surgery (Tab. 2). The pre-surgery questions addressed (1) the reasons for choosing surgery, (2) information sources, and (3) personal emotions and attitudes. After surgery, they were asked about (1) health, (2) nutrition, (3) physical activity, (4) self-image, (5) people’s attitudes, (6) psychological issues, and (7) relationships. Within each sub-category, participants were allowed to speak freely about their experiences. Participants emphasized different issues, but there were some commonalities. The questions were further explained as required, and the replies were followed up by clarifying inquiries.

**Table 1.**
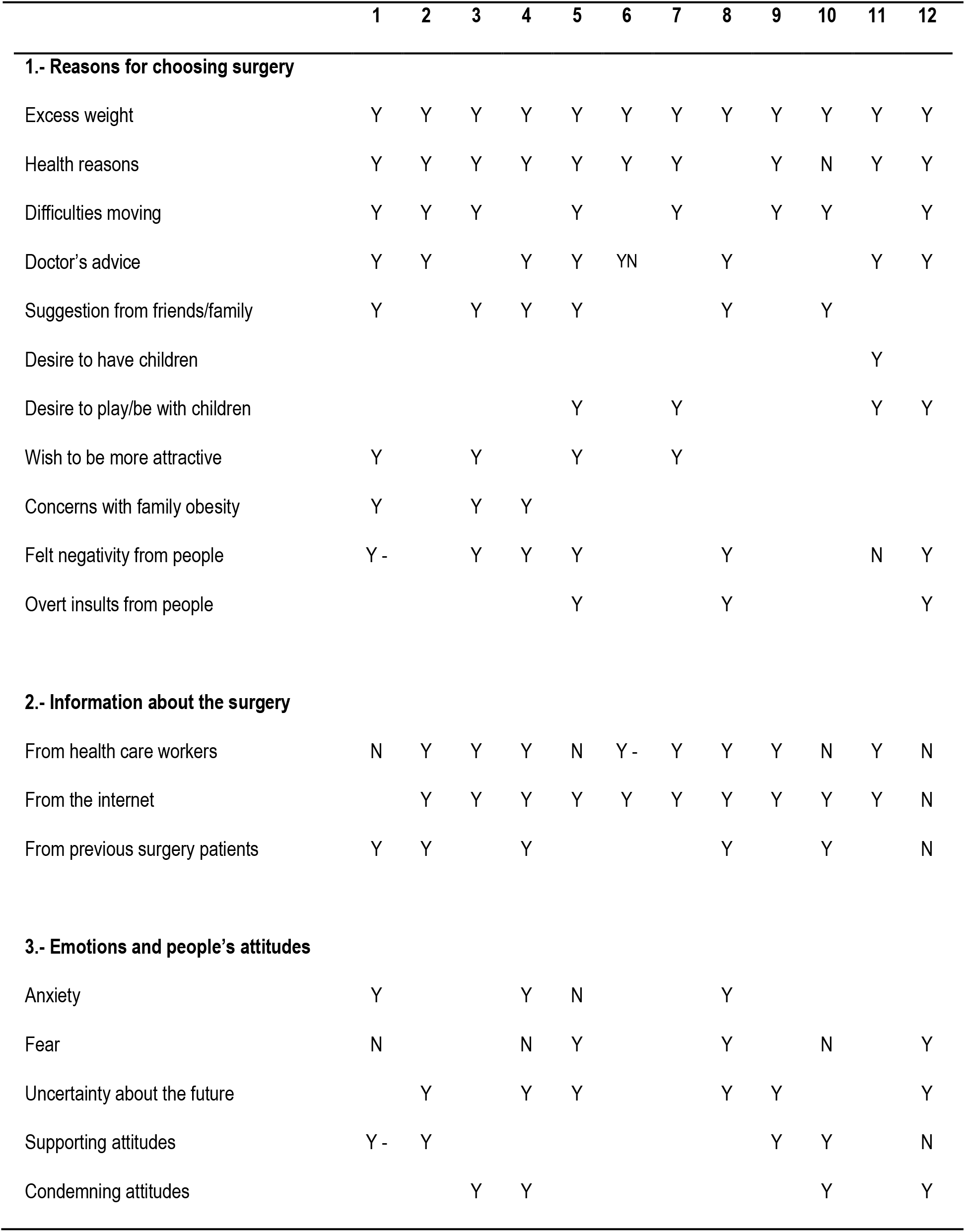
Changes and experiences before surgery. Blank = not mentioned, Y = yes, N = no, - = Yes, but not enough. See the text for more detailed explanations of the categories.

**Table 2.**
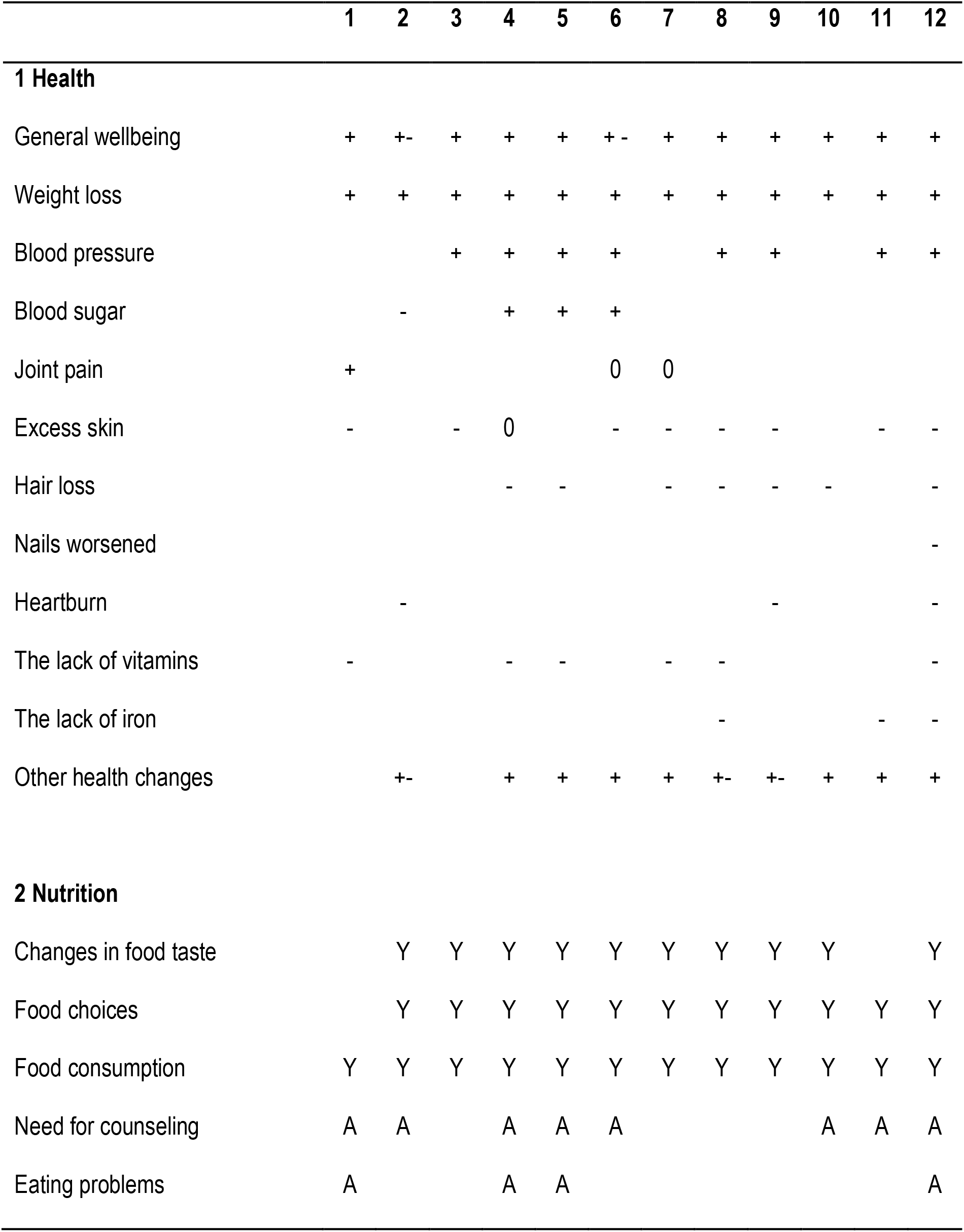
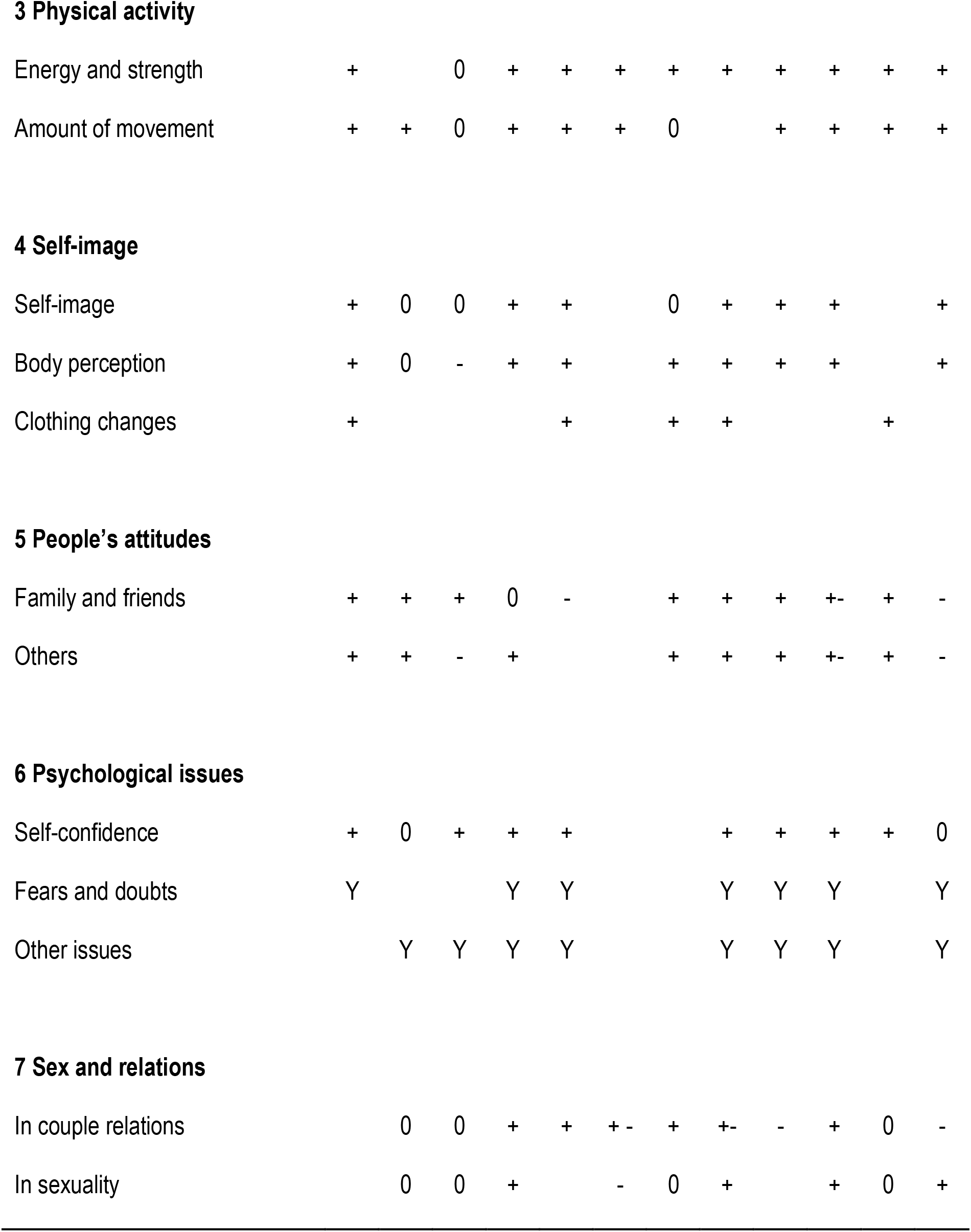
Changes and experiences after surgery. Blank = not mentioned, 0 = no change, Y = changes either positive or negative, + = positive change, - = negative change. A = agree. See the text for further explanations about the categories.

|  | 1 | 2 | 3 | 4 | 5 | 6 | 7 | 8 | 9 | 10 | 11 | 12 |
| --- | --- | --- | --- | --- | --- | --- | --- | --- | --- | --- | --- | --- |
| **1 Health** |  |  |  |  |  |  |  |  |  |  |  |  |
| General wellbeing | + | +- | + | + | + | +- | + | + | + | + | + | + |
| Weight loss | + | + | + | + | + | + | + | + | + | + | + | + |
| Blood pressure |  |  | + | + | + | + |  | + | + |  | + | + |
| Blood sugar |  | - |  | + | + | + |  |  |  |  |  |  |
| Joint pain | + |  |  |  |  | 0 | 0 |  |  |  |  |  |
| Excess skin | - |  | - | 0 |  | - | - | - | - |  | - | - |
| Hair loss |  |  |  | - | - |  | - | - | - | - |  | - |
| Nails worsened |  |  |  |  |  |  |  |  |  |  |  | - |
| Heartburn |  | - |  |  |  |  |  |  | - |  |  | - |
| The lack of vitamins | - |  |  | - | - |  | - | - |  |  |  | - |
| The lack of iron |  |  |  |  |  |  |  | - |  |  | - | - |
| Other health changes |  | +- |  | + | + | + | + | +- | +- | + | + | + |
| **2 Nutrition** |  |  |  |  |  |  |  |  |  |  |  |  |
| Changes in food taste |  | Y | Y | Y | Y | Y | Y | Y | Y | Y |  | Y |
| Food choices |  | Y | Y | Y | Y | Y | Y | Y | Y | Y | Y | Y |
| Food consumption | Y | Y | Y | Y | Y | Y | Y | Y | Y | Y | Y | Y |
| Need for counseling | A | A |  | A | A | A |  |  |  | A | A | A |
| Eating problems | A |  |  | A | A |  |  |  |  |  |  | A |

*Table 2.. cont.***3 Physical activity**
|  |  |  |  |  |  |  |  |  |  |  |  |  |
| --- | --- | --- | --- | --- | --- | --- | --- | --- | --- | --- | --- | --- |
| Energy and strength | + |  | 0 | + | + | + | + | + | + | + | + | + |
| Amount of movement | + | + | 0 | + | + | + | 0 |  | + | + | + | + |
| **4 Self-image** |  |  |  |  |  |  |  |  |  |  |  |  |
| Self-image | + | 0 | 0 | + | + |  | 0 | + | + | + |  | + |
| Body perception | + | 0 | - | + | + |  | + | + | + | + |  | + |
| Clothing changes | + |  |  |  | + |  | + | + |  |  | + |  |
| **5 People's attitudes** |  |  |  |  |  |  |  |  |  |  |  |  |
| Family and friends | + | + | + | 0 | - |  | + | + | + | + - | + | - |
| Others | + | + | - | + |  |  | + | + | + | + - | + | - |
| **6 Psychological issues** |  |  |  |  |  |  |  |  |  |  |  |  |
| Self-confidence | + | 0 | + | + | + |  |  | + | + | + | + | 0 |
| Fears and doubts | Y |  |  | Y | Y |  |  | Y | Y | Y |  | Y |
| Other issues |  | Y | Y | Y | Y |  |  | Y | Y | Y |  | Y |
| **7 Sex and relations** |  |  |  |  |  |  |  |  |  |  |  |  |
| In couple relations |  | 0 | 0 | + | + | + - | + | + - | - | + | 0 | - |
| In sexuality |  | 0 | 0 | + |  | - | 0 | + |  | + | 0 | + |

Interviews were audio-recorded and subsequently transcribed using a speech recognition program^44^. The transcriptions were corrected as required, using the recordings for confirmation.

### Ethical Considerations

Participants were informed about the goal of the research, the fact that it is voluntary to participate, and that they have the right to refuse to answer any questions, and to remove themselves from the study at any stage. All participants agreed to continue until the end. Participants read, filled, and signed an informed consent form. The data were collected anonymously. The research was approved by the Tartu University human research ethics committee (protocol number 294/T-26).

## RESULTS

Of the 12 interviews, nine were in person and three online via Skype; 10 participants were female and 2 were male. The interviews were conducted from August to October, 2019. The shortest one lasted 11 min. and the longest one lasted 32 min.

### Choosing and preparing for surgery

There were 3 general questions: (1) reasons for choosing bariatric surgery, (2) getting information about the procedure, and (3) personal emotions and attitudes of other people before the surgery. Nineteen general topics were identified (Tab. 1).

#### Reasons for choosing bariatric surgery

In all cases, attempts to lose weight via more conventional means, diet and exercise, had not been successful. The two main reasons why people chose to undergo bariatric surgery were because they were “overweight” (technically “obese”), and because of the associated health problems. The main health problems mentioned were high blood pressure, type-2 diabetes, thyroid malfunction, and joint pain. The latter likely contributed to the third main reason: difficulties moving. The one exception not affected by those reasons was more pragmatic and decided to undergo bariatric surgery because of a doctor’s advice and suggestions from family members. In some cases, the surgery had been recommended by a friend or family member who had already undergone bariatric surgery.

Other reasons included the desire to be more sexually attractive, to play or interact more with children or grandchildren, and in one case, to have children in the future. The category “family obesity” refers to the concern expressed by three participants of gaining even more weight in the future because their family members had excess weight that had increased with age. Some participants mentioned that they **felt** negativity from people, and others indicated that they had **actually experienced** overt negativity and insults. One participant had sad memories from high school when she was teased and humiliated.

#### Obtaining information about bariatric surgery

Participants obtained information from doctors, the internet, and other patients. Most participants said that they were sufficiently informed by their doctor, but others disagreed, or stated that others, but not themselves, would benefit from additional information. Other than that, there was a lot of information in the internet forums and particularly from the Facebook group we used to recruit the participants. Some participants indicated that they would have liked to get more specific information from doctors instead of having to look it up on the internet.

#### Personal emotions and other people’s attitudes

Before the surgery, participants felt anxiety and fear. Patients were mostly afraid of dying on the surgery table, complications from the surgery, and dealing with life after the surgery. Participants were also worried about how others would react to their decision to go through the surgery. About the decision to go to surgery, there was support from relatives and friends, but there were also people who were doubtful and hesitant.

Some participants initially had a negative attitude towards surgery because they thought that surgery was the easy route, instead of just eating less and exercising more. People also thought that there might be complications during the surgery and their health might even get worse. Finally, they feared that they would not be able to change their nutrition anyway and would end up gaining back the weight.

### Life after surgery

There were seven general topics: (1) health, (2) nutrition, (3) physical activity, (4) self-image and body perception, (5) people’s attitudes, (6) psychological issues, and (7) relationships. Twenty-nine general themes were identified (Tab. 2).

#### Health

This category included general well-being, weight loss, blood pressure, blood sugar, joint pain, excess skin, hair loss, nail quality, heartburn, lack of vitamins, lack of iron, and other health changes.

Eight people had the surgery without complications and four people had complications. One person had a longer time of surgery and a larger wound because of liver problems. A second person had a leak of the stomach and peritonitis and needed to stay longer in the hospital. The third person had a fever and infection that was cured after a few days in hospital. Finally, a woman found out after surgery that she was pregnant but fortunately, she later gave birth to a healthy baby.

Everyone lost weight. Nearly everyone reported an increase in wellbeing. Nine participants reported that their previous hypertension was alleviated and about half reported blood sugar levels had improved.

Fast weight loss left excess skin but most of the people accepted it as inevitable and dealt with it with suitable underwear. A couple of people removed the excess skin via plastic surgery and a few had planned to do so in the future.

Several participants experienced hair loss 4-6 months after the surgery but it grew back, sometimes even thicker and healthier. Joint pain was mentioned three times. In one case, it decreased. In two cases, it did not change.

There were also several negative consequences. Some people mentioned heartburn and the need for anti-acid medication. Also, there were people who had swings of blood sugar and dumping syndrome. There were also problems with digestion, dryness of skin, vitamin deficiencies (particularly group B vitamins), iron-deficiency, and anemia. A couple of women mentioned changes in their menstrual cycles; in one case, the cycles became more regular, and in the other, the menstruation became longer and more profuse (menorrhagia). Many people mentioned feeling cold and needing warmer clothing.

However, despite some negative changes in health, most people said that they got rid of numerous health concerns or the health problems eased significantly.

#### Nutrition

Participants mostly mentioned a decrease in the amount of food they consumed, along with changes in their taste and food preferences. About half of the people searched for information independently online, changed their diets, tested their suitability, and established new balanced diets. The other half had difficulties from the start about how much and what to eat. At least three people kept eating the same amounts and vomiting afterwards. There were also problems with digestion and dumping syndrome. Many formerly preferred foods lost their appeal, and some foods that were previously unpleasant became enjoyable. Some interviewees mentioned that eating was not pleasurable anymore and they only ate because of the need to eat. People indicated there was insufficient information from health care workers and the materials and instructions they had received.

One person developed an eating disorder. The participant thought about food constantly and then decreased the amount of food consumed to the point giving up food completely and losing too much weight. Eventually, this person sought psychiatric help and recovered.

#### Self-image and body perception

We differentiated between two closely linked concepts: “self-image” (who you are) and “body perception” (how you view your body). There was an increase in self-satisfaction: people were happier, became more social, and felt more attractive. Both because of the weight reduction and the psychological effects, several participants indicated that they had changed their wardrobe. The size of clothing changed, but also the style. Women started wearing more colorful and revealing clothing. If in the past, people had difficulties to find clothing that fit but now they had more options. Women often changed their hair color.

#### People’s attitudes to the surgery

The attitudes of close people were both positive and negative. Most participants’ relatives were positive and supportive. In contrast, several people said that their family members were strongly against the surgery and did not support them in their choice. A couple of people actually hid their decision to have the surgery and revealed it only after the procedure had been completed. The attitude of friends, colleagues and acquaintances was generally positive but there were also negative opinions.

#### Psychological Issues

Most people experienced an increase in self-confidence. They felt more acceptable in society, and more attractive to opposite sex. Women felt more feminine. Both men stated that they were self-confident before the surgery and in that respect did not feel any different. The increase in self-confidence led to greater courage communicating with people, and a desire for new activities and directions in life, for example, to go back to school.

On the other hand, there were also negative effects. The biggest concern was that the weight loss would not be permanent. In some cases, the degree of self-criticism increased and people were still unhappy with their own image. One person developed body dysmorphia, followed by self-injury/cutting, and eventually recovered with psychiatric help. Some people had problems with depression and insomnia. In general, people stated the need to get psychological counseling before and after surgery. They indicated the need for emotional support from professionals when they did not get it from their partner or family.

#### Relationships and sex

For most people who already were in relationships, their sexual relationship improved, but in some cases, earlier problems were exacerbated. Oddly, one reason mentioned for a break-up was that the partner did not like the new slimmer body and he/she preferred the previous lush figure. People found new partners, married and had children. Some participants, including both males, did not report any changes in their relationships or sexuality. Single people, including most single women, found more self-confidence, started communicating and socializing more, and had more sexual relationships.

## DISCUSSION

Obesity is a global epidemic and a major health concern and bariatric surgery is an increasingly common treatment option. Here, we examined people’s experiences before and after bariatric surgery. Experiences **before** surgery were divided into: (1) reasons for choosing surgery, (2) getting surgery information, (3) personal emotions and attitudes. Experiences **after** surgery were grouped into: (1) health, (2) nutrition, (3) physical activity, (4) self-image, (5) people’s attitudes, (6) psychological issues, and (7) relationships.

### Experiences before surgery

**The reasons for choosing bariatric surgery** were unsurprising: people were overweight, had related health problems, had difficulties moving, and hoped for positive changes, such as having a family and looking better. Other researchers report similar findings^45-47^, which can be summarized as primarily medical reasons first and secondarily, psychological and social reasons.

A Brazilian study of 12 women stands apart in that the motivation for bariatric surgery was mostly social: the desire to look better, to be more accepted in society, and to enter the labour market^48^. Cohn *et al*.^49^ also point out that people choosing bariatric surgery desired to improve their professional lives. Participants in our study did mention negative attitudes towards overweight people, but did not mention any such experiences specifically at their place of work. Overt discrimination and negative attitudes towards obese people at the workplace might not be as prevalent in Estonia.

**Information about bariatric surgery** came from medical professionals and the internet. Participants used official medical sites on the internet, but they also indicated that they valued social media groups where people share common experiences. Cohn *et al*.^49^ offered the same conclusion. In our study, most people thought that they had received sufficient information from their physicians, but others disagreed. Whereas attending information meetings in person can also contribute to the decision to undergo bariatric surgery^49^, this factor was not mentioned by the participants of our study, who instead preferred to communicate online.

**Personal emotions and people’s attitudes before surgery** were complex. Family members’ attitudes were bipolar. Most participants’ relatives were supportive but several were against the surgery. Similarly, friends’, colleagues’, and acquaintances’ attitudes were generally positive but there were also negative opinions. There was uncertainty, anxiety, and apprehension about the surgery itself, personal changes afterwards, and reactions from other people. Similar psychological and sociological concerns have been reported recently in the literature^5,49^. Hence, the decision to undergo bariatric surgery, although primarily motivated by health concerns, is also strongly affected by psychological and social factors.

### Experiences after surgery

**Health** improved as expected. In the immediate aftermath, four out of 12 people had post-operative complications, but not after they had left the hospital. Physicians dealing with emergencies of bariatric surgery patients need to be know the type of surgery performed and the common emergencies arising from bariatric surgeries^50^.

Everyone reported weight loss, so the main goal of the surgery was achieved, and several related health problems also decreased. This is not unexpected^51^ and it explains the increasing popularity of bariatric surgeries. However, weight loss and the associated health benefits are highly variable^52^. Skin and hair problems are to be expected due to micronutrient and mineral deficiencies^53^, but in due time, people in our study managed to alter their diets as required. Other studies have reported a decrease in joint pain, particularly in the knee^51^. In our study, joint pain decreased in one case, but did not change in two cases. An explanation might be that due to the previous weight, leg joints were already damaged, perhaps permanently.

Several people mentioned feeling cold, which has not been reported elsewhere, but is not particularly surprising, as a layer of fat is a common thermal insulator for mammals living in cold environments^54^. This discomfort must be weighed against the benefit of becoming less susceptible to heat stress, which might be important in warmer climates but perhaps not so much in Estonia, other than in the sauna. Several other minor health concerns were mentioned, but the net effect was that bariatric surgery had a positive effect, not only weight loss, but also improving related health problems.

**Nutritional changes** dealt mostly with the amount of food consumed, and changes in food preferences and flavors. Similar findings have been reported elsewhere^55^. In our study, about half the people found information on their own and changed their diet as necessary. The other half had difficulties from the start and would have benefited from receiving more information and support from medical personnel^55^.

Another important but not unexpected effect was a deficiency in some micronutrients: iron, zinc, and vitamin B12. Micronutrient and mineral deficiencies can be manifested as readily visible skin problems^53^. Solving this issue is usually a matter of being aware of the possibility and taking the right nutritional supplements^56^. Nutrition support from professionals might be particularly important for some patients^57^, about half in our sample.

After surgery, one participant in our study developed what might be considered the opposite disorder. From its description, it approached anorexia nervosa, even if was not formally diagnosed as such. Oddly, obesity is officially classified as a nutritional disorder, but anorexia nervosa as an eating disorder^58^. However, both obesity and anorexia nervosa share many biological, psychological, and sociological components and might actually be two extreme expressions of the same predispositions^59^. At the population level, the two disorders might even be causally linked^21^.

**Physical activity** increased substantially: moving became easier and more pleasant, the distances became longer, and people had more stamina and strength. This has been reported elsewhere^60^, with the caveat that some improvements decreased after one year^61^. Other studies have reported a decrease in joint pain, particularly knee pain^51^. In our study, one person mentioned a decrease in joint pain decreased in one case, but two people indicated that it had not changed. An explanation might be that due to the previous weight, leg joints were already damaged, perhaps permanently.

**Self-Image and body perception** did not necessarily change, but people generally felt happier, more social, and more attractive. Others have reported cases whereby patients struggled with their self-identity^36^, but in our study, it was not an issue. About half of men are satisfied with the results of their surgeries, compared to only 12% of women^62^. Women in our study reported more changes in clothing style than men did, so changes in women, unlike those in men, had more external manifestations and were more readily visible to society.

**People’s attitudes** after the surgery continued being both positive and negative, just like before the surgery, whether from family, friends, colleagues, or acquaintances. Benson Davies *et al*.^63^ concluded that family support is important to maintain the weight loss up to 6 years after surgery. Perhaps more important than the attitudes towards the surgery itself and the weight loss, attitudes towards the ensuing behavioral and lifestyle changes might be more contentious. Bariatric surgery leads to changes in self-worth, physical activity, psychological state, and life directions, changes that are bound to lead to new and different social interactions.

**Psychological issues** were generally positive, with improvements in self-esteem leading to a more positive attitude in life. However, just because the changes were generally positive does not mean they were easy to manage. Furthermore, there were some problems with depression, addiction, self-injury, and insomnia. Our study’s participants indicated the need for emotional guidance from professionals, particularly when family members were unsupportive. The potential benefits of psychological counseling, both before surgery to prepare patients for the forthcoming changes, and after to help them navigate these changes and identify problems before they develop, have been previously noted^55^. Detailed cost-benefit analyses need to be conducted to determine the degree to which more extensive psychological counseling can become an integral part of the procedure.

**Sex and relationships** generally improved but there were some cases whereby problems couples already had, actually intensified. Some people became more sexually active and others did not report any differences. Single people became more sexually engaged and sexually active. Despite a better sex life, some marriages deteriorated.

We did not have enough male participants to note differences between the sexes. In couples, “sexual function” of bariatric surgery patients and their partners generally improves^64^. However, among single people, “sexual satisfaction” increases in women, but not in men^65^. This is despite that erectile function and testosterone do increase in men after losing weight^66^. This apparent incongruity occurs because “sexual function” is not the same as “sexual satisfaction”. For males, bariatric surgery might improve sexual function, placing patients at par with every other male and still facing the main hurdle: access to women. In contrast, for females, access of males is seldom a problem. Female sexuality is not about the actual act, but rather about receiving attention from men and then choosing to act (or not) as they see fit. It is reasonable to expect that the sexual benefits of losing weight, whether from bariatric surgery or otherwise, will be greater in women than in men.

Future work on the sexuality and relationships of bariatric surgery patients within couples would benefit from comparing them with regular couples, obese couples who underwent no surgery, couples undergoing other comparable life-altering procedures, and of course, cases in which the male, the female, or both partners undergo the procedure.

**Finally, two other issues came up:** the sex ratio of bariatric patients and the cost of the procedure. First, based on the literature and this study, most patients tend are female, and hence, most studies are female biased and might not be directly applicable to males. Obesity is female biased in developing countries, but male biased in developed countries^67^. Medical practitioners ought to do more to address this bias and perhaps attract more male patients^68^. Why are more women than men attracted to the procedure? Of all the benefits that we examined and found in the literature, only one, sex and relationships, seems to benefit women more than men. Is it important enough to explain the effect?

Second, in Estonia, the state covers the costs of bariatric surgery for residents. In countries where the government does not pay, the cost weighs heavily on the decision to undergo the procedure^46^. Whether the procedure is financed individually or by the state, bariatric surgery does reduce the total costs of health care over a person’s lifetime^69^. Public health officials and economists will have to work together and decide the degree to which bariatric surgery will continue to play a role in the battle against obesity^70^.

In conclusion, bariatric surgery leads to large physical changes, which, in turn, have drastic effects on people’s everyday lives, their internal mental states, and their roles in society. Potential patients ought to be better prepared for these changes, have more realistic expectations, and be willing to seek counseling both before and after surgery. Concurrently, medical practitioners ought to be aware of the magnitude of the changes this surgery will bring about in their patients’ lives, and help them to prepare for surgery and to adjust to the changes afterwards.

## Data Availability

All data produced in the present study are available upon reasonable request to the authors

## ACKNOWLEDGEMENTS

We thank the participants in the study.

## AUTHOR CONTRIBUTIONS

Study concept and design: TP and LL; acquisition of data: TP and LL; analysis and interpretation of data: all authors; initial drafting of the manuscript: LL; final writing and critical revision of the manuscript: GAL.

